# Lived experiences of pregnant and new mothers during COVID-19 pandemic: A narrative analysis of YouTube birth stories

**DOI:** 10.1101/2020.12.28.20248958

**Authors:** Kobi V. Ajayi, Idethia S. Harvey, Sonya Panjwani, Inyang Uwak, Whitney Garney, Robin L. Page

## Abstract

**Introduction:** The COVID-19 pandemic has brought on unprecedented changes, not only to our daily lives but also to our healthcare system. The pandemic has particularly impacted pregnant women that must give birth with tight restrictions and significant uncertainties. Birth stories have frequently been used as a way for women to describe their experiences with the birthing process. In this uncertain time, birth stories can provide valuable insight into how pregnancy and birth stressors during a pandemic can impact the patient’s overall experience. This study sought to describe and understand pregnant and new mothers’ lived experiences during the COVID-19 pandemic.

**Methods:** Researchers extracted relevant YouTube birth stories using predetermined search terms and inclusion criteria. The mothers’ birth stories were narrated in their second or third trimester or those who had recently given birth during the study period. Birth stories were analyzed using an inductive and deductive approach to capture different aspects of the birthing experience.

**Results:** Overall, eighty-three birth stories were analyzed. Within these birth stories, four broad themes and twelve subthemes emerged. Key themes included a sense of loss, hospital experiences, experiences with healthcare providers, and unique experiences during birth and postpartum. The birth stories revealed negative and positive birth experiences. Particularly, mothers were frustrated with constantly changing policies within the healthcare setting that negatively affected their birthing experience. On the other hand, support from healthcare professionals, having their partners in the delivery room, and having a positive mindset was instrumental in having a positive birth experience.

**Conclusion:** Results from this study provided a detailed description of women’s lived experience with giving birth during the COVID-19 pandemic. Healthcare providers need to provide clear communication and compassionate patient-centered care to relieve women’s anxiety about uncertain and unpredictable policy as the pandemic continues to evolve.

## Introduction

In our novel coronavirus (COVID-19) world, can birth stories help guide clinical practice to achieve a positive birth experience?

Empirical studies suggest that pregnant women with COVID-19 are at an increased risk of severe illness and adverse pregnancy outcomes such as preterm birth.^1^ Studies have also reported elevated levels of stress and anxiety among pregnant women due to the pandemic.^2^ Although the pandemic-related stressors (e.g., social isolation, quarantine, face mask, misinformation, and uncertainties) are not unique to this population, changes to hospital and provider policies (e.g., suspension or cancellation of in-person antenatal care and policies restricting visitors during labor and delivery) may lead to adverse birth experiences and poor mental health outcomes.^3,4^ Studies posit that a negative birth experience, having a sudden change in birth plans, and lack of support during birth are associated with post-traumatic stress disorder (PTSD).^5^ Moreover, mental health problems during pregnancy are associated with lifelong negative consequences on the mother and fetus.^5^ Given these concerns, identifying ways to achieve positive birth experiences as the pandemic evolves is paramount.

Scholars assert that insights from birth stories can lead to a therapeutic relationship between pregnant women and their healthcare providers, which studies suggest can guide clinical practice, design care protocols, and facilitate a positive birth experience.^6^ Because “birth stories are personal narratives grounded in the pivotal life experience of giving birth,^7^” this study is grounded in the underpinnings of the narrative interpretative framework. According to Clandinin and Connolly in Creswell & Poth, “narrative inquiry is stories lived and told” ^8^ and can be collected as oral or documented collections^8^. Specifically, the narrative analysis explores an individual’s lived experiences within the social, familial, linguistic, and institutional context that shapes the experience^8^. Evidence shows that narrative medicine is effective in shaping health outcomes and policies across several health domains.^9^ Put simply, narrative medicine improves communication between patients and physicians, quality of care, and health outcomes.^10^ Moreover, the constellation of clinical practices and narrative medicine provides intellectual clarity to care practices, offers rich data to understand essential aspects of the childbearing experience, and improves childbearing women and their families’ care.^6,7,11^ In the pandemic context, birth stories may provide a mechanism that can effectively point out potential problems with COVID-19 obstetric care protocols, evaluate existing hospital policies, and examine the unintended consequences of these policies. Therefore, birth stories can inform the clinician’s understanding of women’s experiences that may facilitate a positive birth experience.

Birth stories are told through several mediums, including the internet. The internet has a profound effect on decision making across a broad range of health topics, including pregnancy and childbirth.^12–14^ Internet platforms such as YouTube offer free video-sharing capabilities that attract millions of viewers and provide a sense of community among users (www.YouTube.com). For these reasons and more (e.g., the ability to learn from views), YouTube has become a one-stop-shop for pregnant women who often share their birth stories.^15,16^ Due to YouTube’s broad reach, video-sharing capabilities, and accessibility, this study draws its data from birth stories shared on YouTube. Analyzing videos on YouTube enables researchers to conduct in-depth qualitative analysis through user-generated birth stories.

The primary purpose of this study is to provide a detailed description of new mothers’ lived experiences during childbirth in the context of COVID-19 and assess what mothers feel their health care provider should have done differently in response to COVID-19. We also aim to use their stories and perspectives to provide answers to the burgeoning questions expectant mothers might have. Results from this study will guide health care providers, policymakers, and researchers on how to incorporate patients’ voices to guide a positive birth experience.

## Methods

### YouTube search

Data was extracted from YouTube site using the following keywords “coronavirus,” AND “pregnancy,” AND “birth story” on September 9, 2020. The extracted videos were sorted using the default YouTube algorithm (Sort by-relevance). Additional parameters included (Upload date - year and Type - video).

### Inclusion and exclusion criteria

Videos were included in this narrative analysis if they were in English, first-person account of the birth story, and narrated by women (with or without their partners) who were either pregnant in their second or third trimester or new mothers who gave birth within the study period. We excluded videos from news agencies, hospitals or health-related organizations, healthcare professionals, duplicate videos in whole or in part, or those from commercial or promotional sources as seen in Figure 1.

**Figure 1:**
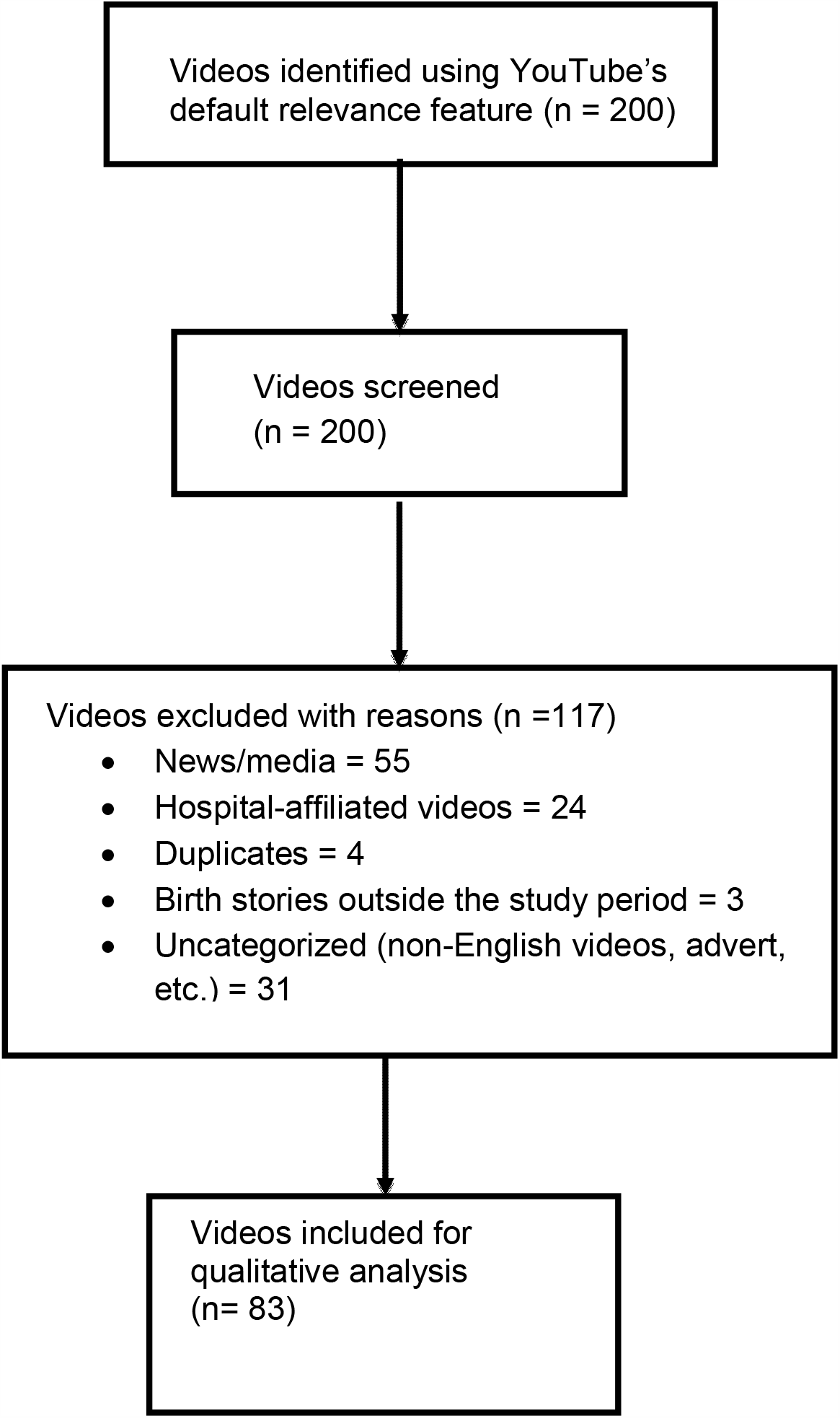
Flow diagram of birth stories included for narrative analysis.

### Data extraction and synthesis

Given that internet users seldom go beyond the first pages of any search,^12^ we restricted our initial sample to the first 10 pages/200 videos.^17^ Moreover, results from a literature review of YouTube videos found that of the 37 studies included for review, 30 studies restricted their search to a limited number of pages for analyses.^14^

After watching the videos extensively and in full, we extracted and recorded the video URL, titles, length, and dates. We also utilized an inductive and deductive coding strategy to develop the codebook. Next, the researchers conducted two iterations of coding. First, open coding was used to create labels. The second stage of coding was done to identify the relevant categories and thematize the data. To establish scientific rigor and inter-rater reliability, two researchers independently and systematically coded 5% of the included videos by selecting every odd number. The raters agreed on 100% of the extracted videos. After that, one of the team members then coded the remainder videos following the established guidelines. This approach allowed for consistency throughout the coding process.

### Ethics

The study was exempted from ethical approval because YouTube videos are publicly available data.

## Results

### Study characteristics

We analyzed eighty-three birth stories. The majority of the birth stories were from the United States (72%), including women in the postpartum period (77%), were singleton deliveries (86%), and were vaginal births (63%). The videos ranged from 2 minutes to 1:02:34. Detailed information about the video characteristics is found in Table [1]. The narrative analysis revealed four broad themes and twelve sub-themes.

**Table 1.** Characteristics of birth stories on YouTube between January and September 2020. (n = 83)

| Characteristics | Values |
| --- | --- |
| **Country, n (%)** |  |
| United States | 60 (72.2) |
| United Kingdom | 8 (9.63) |
| Canada | 6 (7.22) |
| Australia | 2 (2.4) |
| Spain | 1 (1.2) |
| Jamaica | 1 (1.2) |
| Philippines | 1 (1.2) |
| Switzerland | 1 (1.2) |
| Not reported <sup>a</sup> | 3 (2.31) |
| **Period birth story was told, n (%)** |  |
| Pregnancy | 4 (4.81) |
| Postpartum period | 67 (80.7) |
| Labor/delivery | 12 (14.4) |
| **Type of delivery, n (%)** |  |
| Vaginal | 55 (66.2) |
| Cesarean delivery | 22 (26.5) |
| Not reported <sup>a</sup> | 6 (7.22) |
| **Type of birth, n (%)** |  |
| Singleton | 76 (91.5) |
| Twin/triplet | 4 (4.81) |
| Not reported <sup>a</sup> | 2 (2.4) |
| **Parity, n (%)** |  |
| Primiparous | 41 (49.3) |
| Multiparous | 31 (37.3) |
| Not reported <sup>a</sup> | 11 (13.2) |
| **Delivery term** |  |
| Full-term <sup>b</sup> | 72 (86.7) |
| Preterm <sup>c</sup> | 5 (6.02) |
| Not reported <sup>a</sup> | 6 (7.22) |

a Videos did not explicitly reveal the information for these categories.
b defined as birth > 37 weeks. c defined as births <37

### Themes

#### Sense of Loss

An overall sense of loss of the pregnancy and birthing experience under normal conditions (i.e., the ability to have a baby shower or have family present after giving birth) was a common theme conveyed within the videos. As one woman expressed, “Obviously, being pregnant, one of the things you do is have baby showers. And I had three of them scheduled, and they were all going to be in the month of April. […] I remember asking my doctor what she thought about doing these baby showers. And she was like, ‘you’re probably not going to be able to do those.’ So, at that point, we had to make the decision to cancel our April baby showers, which was a huge bummer. And being so pregnant and emotional, I was really upset about it.”

Similarly, the inability to have family members meet the baby after birth also created a sense of loss: “Because of COVID-19, no one other than the spouse was permitted at the hospital with me, so [husband’s] grandparents had to meet him via FaceTime. On the day [baby] was released from the hospital, Ohio was placed on lockdown, and everyone was mandated to stay home. For that reason, [baby] has not yet met his grandparents, aunts or uncles, nor close friends.”

Few women experienced a sense of loss because visitors, including spouses/partners, were not allowed during labor and/or delivery. “Triage says, ‘okay, we want to keep you, but by yourself.’ […] When they said that, I lost it. I started crying. […] I was so upset. It was like a slap in the face. This is my time, you know, you see those videos […], and you all are telling me I have to be alone?”

#### Hospital Experience

Due to the pandemic, hospitals modified their check-in process. Most women had to either call in when they reached the hospital or enter through a specific entrance: “As soon as we walk from the garage to the door of the hospital, we are greeted with a nurse covered from head to toe in protective gear. They stop us to take our temperature, check our eyes, and tell us to go to the next station. There, we go through a questionnaire and meet a police officer who then gives us wristbands to admit us access to the hospital, and he escorts us to the elevators to triage.”

Hospitals also implemented specific procedures to test patients as they walked in: “Going in, I was told obviously we have to wear a mask. And I was going to get the COVID test and not my husband or your partner because they said, if I have it, the chances are he has it. So, we walk through the hospital, and we get a temperature test and a classic questionnaire. And I was like, well, maybe that was the only test that we need, which is great because I was nervous about getting the thing stuck up my nose.”

Some obstetricians (OB) shared information with their patient about COVID procedures during their last appointments, but since the situation was constantly changing, there were some inconsistencies: “at the OB, they had told us that they were giving people the option to test for COVID, and then when we got to the triage, they were like we’re testing for COVID. It wasn’t an option.”

Masks were mandatory in most cases, which created a challenge for women: “As soon as we walked in, we were given a mask. And we had to wear a mask the entire time that we were there. The only exception was when we were in our recovery room, and it was just me and [husband]. But every time a nurse would come in, we had to put the mask on. And I had to wear the mask during the C-section, which was, oh my gosh, that was a mess, honestly.”

#### Experience with Healthcare Provider

Women in this study revealed that having clear communication and discussing their care options with their provider was soothing. One woman expressed,

“… I got to ask all of my questions […] the nurses came in and introduced themselves…then the anesthesiologist came in and explained everything from start to finish, it brought so much peace of mind having him explain everything and talk me through everything […], and it made me feel just so much more calm going into it [Cesarean delivery].”

Women who gave birth during the pandemic expressed that the support they received during labor/delivery was instrumental in having a positive birth experience. One woman discussing her experience while pushing said, “… it was a beautiful experience because the Matrona, which is the nurse she was like next to the doctor just checking on me. There was a moment while I was pushing, she like got on top of me like to help push over my belly and she was like really comforting, she told me she was going to lower the light so that I can feel more calm and it was just a beautiful experience, and I’m so glad.”

Yet, another mom who was sad because the hospital did not allow a photographer into the delivery room because of the COVID-19 virus noted how her nurses helped her take all the pictures during delivery, which happened to be the only picture with memories of the birth summed it perfectly. “I feel like the nurses and doctors have just gone like above and beyond during this time to make people still feel cared for and feel at ease during this really scary time.”

#### Overall Birth Experience

##### Positive COVID-19 cases vs. non-positive COVID-19 cases

Mothers who tested positive for the COVID-19 virus were unsatisfied with the care they received. One mother who had four COVID-19 tests after subsequent tests were negative following an initial positive test expressed her frustration with the care she received. Her care team told her, “…we know you have three negatives back, but we will still treat you like you have it. The nurses were like they didn’t want to spend a lot of time in the room […], and the lactation nurses declined to come to see me.”

##### Hospital birth vs. home birth

Due to the pandemic, mothers opted for home birth rather than birthing in the hospital to prevent contracting the virus. One mother who has never had a home birth said, “…this coronavirus chaotic time, I had […] fears. I had to last-minute change my birth plans to help me feel comfortable in giving birth.”

##### Bonding with partner

Mothers expressed that hospital policies limiting visitors to one partner made them bond with their partners and baby better both at the hospital and home. As stated by one woman, “…I could also see a lot of positive things, so if any of you guys are going through that [not having family members around] instead of looking at the negative side…, look at it as a really strong bonding moment with your husband…, it’s actually truly beautiful to be able to share that with your partners.”

##### Healthy baby

Despite the restrictive hospital policies, changes to birth plans, and other losses during pregnancy because of the coronavirus, women stated that having a healthy baby diffused the negative feelings of loss from the pandemic. “The main thing was we wanted to have a healthy baby, and all of the things that I missed out on all the craziness that was happening like none of it mattered when it just came down to like the point of all this is for us to just have a baby. All these other things like baby showers and photos and trips like none of that mattered in the midst of just like having a baby.”

##### Mentally prepared

Moms maintained a positive mindset and were mentally prepared for the worst outcome possible (e.g., birthing alone) so that they were able to stay strong and maximize their birth experience. They saw the situation as a way to affirm strength and power over their minds and bodies.

A mom who delivered alone and spent two days at the hospital was sad because her husband had to watch their older child at home, said: “…, so I was already kind of mentally preparing myself, and I was telling myself like you are strong, you’ve got this…you giving birth alone is not the end of the world it’s not the biggest deal…you are going to be really proud of yourself…so I kept telling myself things like that.”

##### Faith

Faith or belief in God or a higher being can provide a source of strength and support for women during childbearing.^18^ In the context of the pandemic, a person’s faith may play a substantial role in enhancing coping skills to mitigate stress and worries that come from the pandemic’s sequelae. Such narratives were seen among mothers in this study.

One mother mentioned how God helped made her nurse change the date of her last ultrasound visit. She said, “God is just so good because anything could have happened. He (God) works everything out, so I’m just so grateful that He did what He did and everything worked out.” In support of her belief, her partner also referenced the role God played in ensuring he could give her emotional support during delivery.

##### Technology

The COVID-19 birthed a new landscape for telemedicine. In this study, mothers resorted to video calling family members using their electronic devices in response to the restriction COVID-19 hospital policies.

Moreover, mothers with babies in the neonatal intensive care unit (NICU) thought that hospital applications were helpful to help them keep in touch with their babies. One mother with a preterm baby expressed her frustration of being unable to visit her baby in the NICU because of the COVID-19 virus said, “What I do like that my hospital do was they Face Timed me. They had like an iPad there, and they will call me and update me and show me, my baby.”

##### Self-advocating

During the pandemic, mothers who gave birth noted that they self-advocated for themselves because they felt their care team was not paying attention to their needs. One mother with twins who had gone in for her routine stress test alone because of the hospital rules had a chilling experience. “I live 35 minutes away from the hospital, so it’s like, why are they sending you home? That doesn’t make any sense […] I kept advocating like hey do you want to keep me […] even my sister called them to talk to them, and they were like no.” She also said, “stay home because of COVID-19 or go to the hospital because you know your baby is coming.”

##### Information and decision about care

A woman who gave birth alone expressed her frustration with her care team and titled her story “Traumatic Birth Story During Pandemic!” She said, “…I don’t know what to think, you know I’m asking like can someone just let me know what is going on because they’re not telling me anything like they’re talking amongst themselves, but they’re not telling me anything…”

The right to access information enables women to make informed decisions about their care. Moreover, “respectful maternity care requires effective communication between patients and providers.”^4^ As such, in the context of the pandemic, upholding clear communication with women is essential to promoting respectful maternity care, which in turn can improve birth experiences.^19^

##### Postpartum adjustment

Mothers noted that they struggled to adjust to the postpartum period. One mother said, “…honestly, during the first month of having a baby, I was topless for most of the day and those first couple weeks I definitely had like the baby blues, and I felt like I just cried all the time.” Another mom, after birth, said, “…I was just so emotionless, I just didn’t have, I wasn’t feeling anything […], I was just really numb […], and I pretty much didn’t sleep, I sat in the toilet expressing colostrum and feeling sorry for myself.”

##### Maternal health conditions

Although some mothers knew about their pre-existing conditions, others who had seemingly healthy pregnancies were shocked to be notified of a potentially threatening health condition during their routine doctor’s visit or the last ultrasound. For example, one mother whose partner resisted the hospital policies to be with her during the ultrasound visit was surprised by the revelation of the ultrasound that she had a very low amniotic fluid and was thus induced and scheduled for an emergency delivery. She noted, “… I was supposed to be here just for an ultrasound, and I’m just here, and I’m not going home.”

##### Child health conditions

Mothers who had an adverse birth outcome noted how hospital policies made their birthing experience particularly difficult. One mother in particular, who had preeclampsia, seizures, passed out for about 20 minutes immediately after birth and had her baby in the NICU because he could not breathe after birth, recounted her struggles. She said, “…I wasn’t able to be in the NICU with my husband […], the hard thing about the NICU that was really hard for us that as soon as I went in to go see baby [name], partner [name] had then lost his right to go see him [baby] due to COVID.”

Another mom whose baby had a cleft palate and Parry-Romberg syndrome^1^ said, “because of COVID-19 guidelines, the hospital basically said to us that only one parent per 24 hours could visit the baby [name]. I don’t understand how you could not see your baby and how you could choose which parents [are] more valuable and which parents should go.”

## Discussion

The “stay at home” orders due to COVID-19 have created difficulties for women seeking prenatal care around the globe. This qualitative study among recently pregnant and postpartum women described their experiences during pregnancy and delivery during the COVID-19 pandemic. The new mothers’ video described how the coronavirus pandemic changed the delivery procedures and how they interact with their family and health care providers. They expressed a sense of loss, their interactions with healthcare providers, unexpected positive experiences, and adverse outcomes during the delivery. The mothers describe a broad array of emotions as they navigate the pandemic due to the anxiety about the risks that occurred during delivery that may impact their babies’ health.^20^

In the birth stories we analyzed, mothers had to advocate for care, and the delivery team did not provide effective communication during the delivery process. Lack of information during pregnancy and delivery increases anxiety and fears among ‘newly-minted’ moms.^21,22^ Childbirth is often characterized by uncertainty for everyone; however, the COVID-19 pandemic has increased anxiety and fear among pregnant mothers.^21,22^ The lack of communication from healthcare providers, rapidly changing healthcare messages, and the consistently evolving medical care policies (i.e., telehealth)^23^ about what to do during pregnancy increase the uncertainty among birth mothers. ^4,22^ Our findings confirm that pre-and postpartum mothers need increased support and assurance by health professionals during pregnancy, delivery, and the postpartum period.^24^

Due to the pandemic, daily routines, social life, leisure activities, and travel restrictions have impacted how “soon-to-be” parents meet their families through social distance and self-isolation practices.^21^ Social isolation can result in loneliness and reduced social interactions, causing pregnant women to experience a complicated process. It is known that social support is necessary to increase resilience in times of crisis, and poor social support is associated with negative psychological consequences such as feeling lonely quarantine in pandemics.^21^ Similar to previous studies, we found that social support from the pregnant woman’s partner, mother, other family members, midwives, and peers in the perinatal period is essential in reducing stress, improving coping skills, preventing depression, and adapting to new roles as a mother during pre-and postpartum.^25^

This study, however, is not without limitations. First, utilizing YouTube birth stories provides limited sociodemographic or other background information of the participants that may have helped provide rich insights into our findings. Our sample may not reflect women’s viewpoints with low socioeconomic status or rural areas as studies show a substantial digital divide between high versus low socioeconomic groups.^26^ Lastly, as with every cross-sectional study, our study only provides a snapshot of the women’s lives that may not reflect other variables that may impact their overall birth experiences.

Despite these limitations, our study provides important contributions from a global perspective to the limited knowledge of the pandemic’s impact on pregnant and new moms.

## Conclusion

This study shows that women experiencing pregnancy and childbirth during the COVID-19 pandemic face new challenges, in addition to the usual stressors of being pregnant and becoming a new mother. Midwives are uniquely suited to support women and their families throughout this process.

Compassionate, patient-centered communication that provides clear, current information is of paramount importance to allay a woman’s anxiety about uncertain and unpredictable policy changes regarding COVID-19 testing and visitor restrictions. Midwives can foster women’s sense of independence and strength by empowering them to participate in decisions regarding their care and find inner strength through their spiritual beliefs.

Women may be particularly vulnerable during the postpartum phase if they lack access to usual social support sources and face isolation. Providers should consider more frequent and earlier contact with new mothers in the days and weeks after giving birth. Screening for depression and anxiety is essential to identify women who need referrals for behavioral health providers’ treatment. The postpartum is also a period when women’s voices should be heard - allowing them the time and space to share their birth stories. Whether as a YouTube video, blog, virtual support group, or more private journal entry, women should be encouraged to share with others about the life-transforming journey of motherhood.

## Data Availability

Data utilized for this study is publicly available.

https://www.youtube.com/

## Footnotes

1 According to the National Institute of Neurological Disorders and Stroke “Parry-Romberg syndrome is a rare disorder characterized by slowly progressive deterioration (atrophy) of the skin and soft tissues of half of the face (hemifacial atrophy), usually the left side.” Parry-Romberg Syndrome Information Page | National Institute of Neurological Disorders and Stroke (nih.gov)

## Notes

### Competing Interest Statement

The authors have declared no competing interest.

